# Atopic dermatitis remission and recurrence are preceded by changes in ex-tissue-resident memory T cell abundance

**DOI:** 10.64898/2026.09.02.26362057

**Authors:** Treasa Jiang, April R. Foster, Anna V. Pournara, Lloyd Steele, Madiha Shabbir, Emily Smith, Kim Evans, Tom Baxter, Kyle Diddams, Lily Wright, Victoria Rowe, Benjamin Rumney, David Baudry, Anna Rose, Phillip Morgan, Rughashiny Maniam, Kris Stewart, John Y.W. Lee, Aya Balbaa, Haerin Jang, Megan K. Levings, Richard Woolf, Andrew E. Pink, Vijaya B.M. Shanmugiah, Muzlifah Haniffa, Catherine H. Smith, Francesca Capon, Satveer K. Mahil

## Abstract

While targeted therapeutics have transformed the treatment of inflammatory skin disorders, symptoms typically return upon drug withdrawal, reflecting the relapsing remitting trajectory of these conditions. The role of tissue-resident memory T (Trm) cells in recurrence is well established, but the contribution of other memory populations is poorly understood. To address this question, we investigated the immune changes underlying drug-induced remission and recurrence in atopic dermatitis (AD). We used single-cell multi-omics to profile serial blood samples (∼1.3M cells; 5 timepoints over 24 weeks) and skin biopsies (spatial transcriptomics at baseline and week 12) from patients receiving an IL4Rα inhibitor. We found that drug-induced AD remission was preceded by an expansion of memory regulatory T cells in blood and skin. These shifts were accompanied by a rapid decrease in the abundance of blood CD103+/CD4+ ex-Trm cells recirculating from skin. Notably, the reduced frequency of ex-Trm cells was apparent after 3 days of treatment, well in advance of clinical remission. The analysis of independent patient cohorts confirmed this and showed that the decline in ex-Trm cell abundance was reversed upon disease recurrence. Thus, we uncovered early shifts in circulating memory T cells, which anticipate the resolution and return of skin inflammation in AD.

## INTRODUCTION

Chronic, immune-mediated skin disorders are characterized by unpredictable, relapsing and remitting disease flares, which have a substantial impact on quality of life ^1^. In this context, clinical recurrence is driven by inflammatory memory, with skin-resident memory T (Trm) cells playing a key role in the return of symptoms ^2^. Interestingly, at least a proportion of Trm cells leave the skin and recirculate in the bloodstream ^3,4^. These ex-Trm cells then re-differentiate into effector and memory T cells with proinflammatory phenotypes ^5^. Despite this pathogenic potential, it is unclear whether the mobilization of ex-Trm cells contributes to disease flares. Likewise, the role that memory regulatory T (Treg) cells play in remission is not fully understood, as patient cells are often dysfunctional ^6^. Thus, we only have a limited understanding of the immune memory shifts that drive the resolution and recurrence of chronic skin inflammation.

*In-vitro* systems lack the complexity required to address this research gap, while animal models cannot recapitulate the relapsing remitting course of human disease. In contrast, longitudinal studies of patient cohorts following commencement and withdrawal of drug treatment offer unique opportunities to investigate disease remission and recurrence.

Here, we focused on atopic dermatitis (AD), a chronic inflammatory skin disorder, affecting 20% of children and 10% of adults globally ^7^. AD is an archetypal Th2-mediated disease, where patient outcomes have been revolutionized by therapies targeting Th2 cytokines. This success is underscored by the marked efficacy of dupilumab, a monoclonal antibody therapy that inhibits IL-4 and IL-13 activity, by blocking the shared receptor subunit IL4Rα ^8^.

While AD symptoms primarily affect the skin, lymphocytes recruited from the bloodstream play an important pathogenic role ^7^. This warrants an integrated analysis of circulating and tissue-resident populations during the resolution of inflammation. Such studies have, however, been lacking, as published investigations ^9–14^ focused on single compartments and/or used low-resolution methods. Moreover, early time points were typically omitted from longitudinal studies and recurrence was not investigated.

To address these issues, we used single-cell multi-omics to longitudinally characterize circulating immune populations in AD patients receiving or discontinuing IL4Rα blockade over a series of time points. We then followed up key changes in skin, using spatial transcriptomics data obtained at baseline and 12 weeks post-treatment ^15^.

We demonstrate that the abundance of circulating and tissue-resident memory Treg cells increased during the resolution of inflammation. While the abundance of skin Trm cells was unchanged, we found that clinical remission was preceded by a rapid reduction in the frequency of circulating CD4+ ex-Trm cells. This change was already detectable after 3 days of IL4Rα blockade. It was also reversed upon disease recurrence, when treatment was discontinued by individuals who had achieved durable clinical remission. Thus, our results suggest that early shifts in circulating CD4+ ex-Trm cells correlate with disease severity and the transition between remission and recurrence

## RESULTS

### Single-cell analysis of 35 AD cases discriminates circulating immune subsets at high resolution

To identify the circulating immune cells contributing to drug-induced remission, we profiled single-cell transcriptomes, protein surface expression and T cell receptor (TCR) repertoires, in 35 AD patients who had received an IL4Rα inhibitor (dupilumab) for 24 weeks and achieved a clinical response (table S1; fig. S1a). All participants donated blood for Cellular Indexing of Transcriptome and Epitopes by sequencing (CITE-seq) and single-cell TCR-seq on day 0, day 14 and week 12 of treatment, with 23 individuals providing additional samples on day 3 and week 24 (Fig. 1a).

**Fig. 1.**
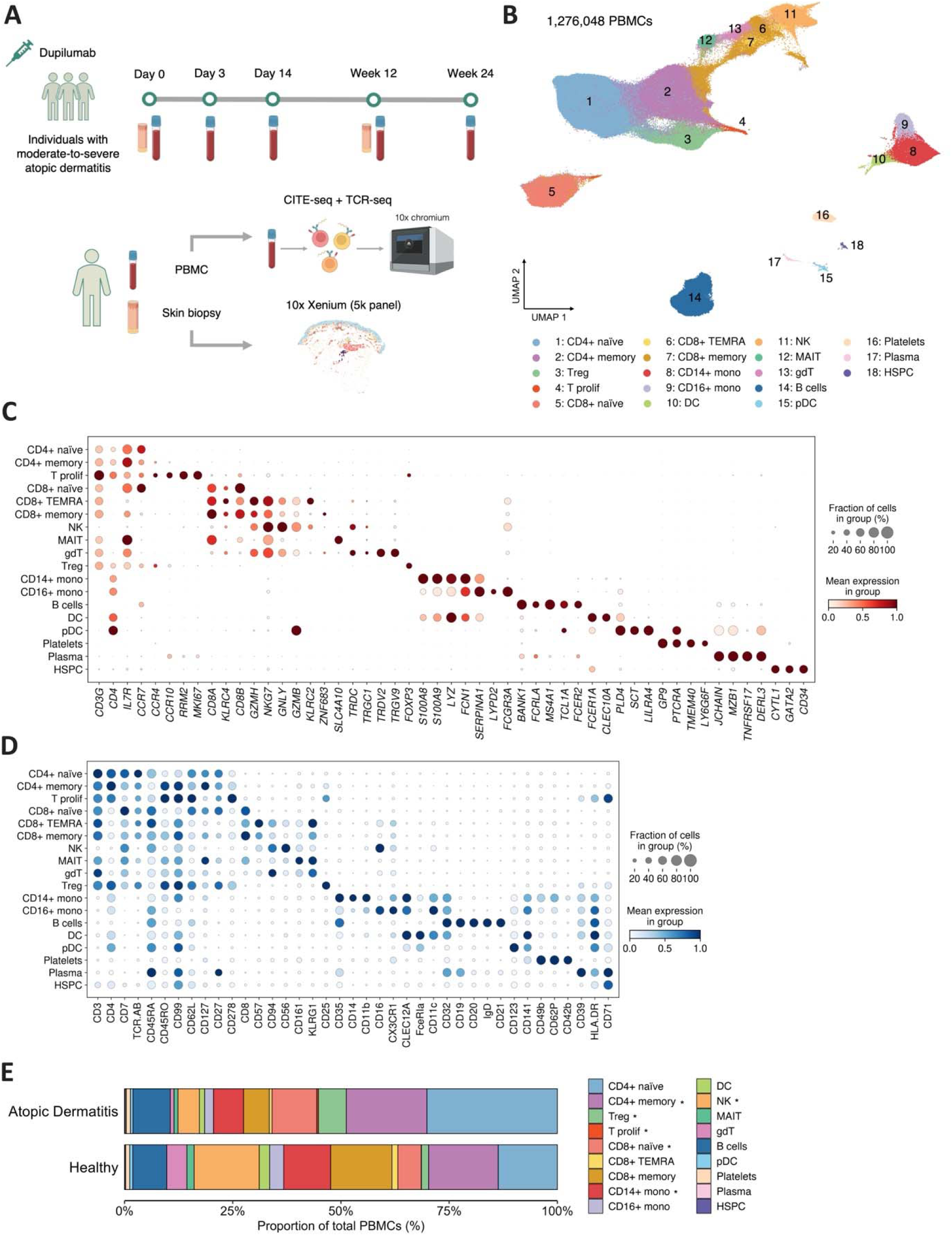
Longitudinal single-cell analysis of drug-induced remission in AD. **A)** Overview of study design, showing the timepoints when blood and skin samples were collected. All 35 participants donated blood on day 0, day 14 and week 12 of treatment, with 23 individuals providing additional samples on day 3 and week 24. Skin biopsies were obtained from 9 participants on day 0 (lesional and non-lesional skin) and week 12 (resolved skin). **B)** UMAP visualization of 1,276,048 PBMCs that passed quality control filtering. **C-D)** Dot plots of CITE-seq RNA (C) and protein (D) readouts, showing the markers used to annotate broad cell types. **E)** Stacked bar plot showing the abundance of broad cell populations in AD cases (baseline values) and healthy controls. *FDR<0.05 (scCODA implementation of differential compositional analysis; CD4+ naïve as reference cell type). DC, dendritic cells; gdT, γδ T cells; HSPC, hematopoietic and progenitor stem cells; MAIT, mucosal associated invariant T cells; Mono, monocyte; NK, natural killer cells; pDC, plasmacytoid dendritic cells; TEMRA, Terminally Differentiated Effector Memory T cells; T prolif, proliferating T cells.

After data quality control, we obtained single-cell profiles for 141 samples and ∼1.3M peripheral blood mononuclear cells (PBMCs) (Fig. 1b and fig. S2a-d). Initial cell clustering and annotation identified 18 leukocyte populations. These included T cells (nine subsets), B cells (two subsets), natural killer cells (NK), monocytes (two subsets), myeloid and plasmacytoid dendritic cells (DC), platelets and hematopoietic stem and progenitor cells (Fig. 1c-d). Interestingly, the integration of a control resource (23 healthy individuals ^16^; fig. S2e) revealed that the T cell compartment was markedly expanded in AD patients compared to healthy donors. This increase mainly affected CD4+ T cell subsets, including CD4+ memory, Treg cells and proliferating T cells. Conversely, and in keeping with published findings ^17,18^, NK cell abundance was markedly reduced among affected individuals (Fig. 1e).

To investigate the resolution of inflammation at higher resolution, we systematically subclustered the main immune compartments. We annotated 13 CD4+ and 11 CD8+ T cell populations. This uncovered multiple naïve, T helper and regulatory subsets in the CD4 compartment (Fig. 2a-c), as well as naïve, memory and exhausted populations among CD8+ cells (fig. S3a-c). We also identified seven B cell clusters, including naïve, transitional and memory populations, plasma cells and plasmablasts (fig. S4a-c). We defined six natural killer cell clusters, corresponding to subsets previously defined by single-cell multi-omics ^19^ (fig. S5a-c). Finally, we observed eight myeloid/plasmacytoid cell clusters, including three monocyte and three DC subsets, a plasmacytoid dendritic cell cluster and a rare but well characterized population of *AXL*+ DCs ^20^ (fig. S6a-c). Thus, the sub-clustering of our dataset enabled us to discriminate 45 leukocyte populations and replicate the findings of other high-dimensional immune phenotyping studies ^16,19,20^.

**Fig. 2.**
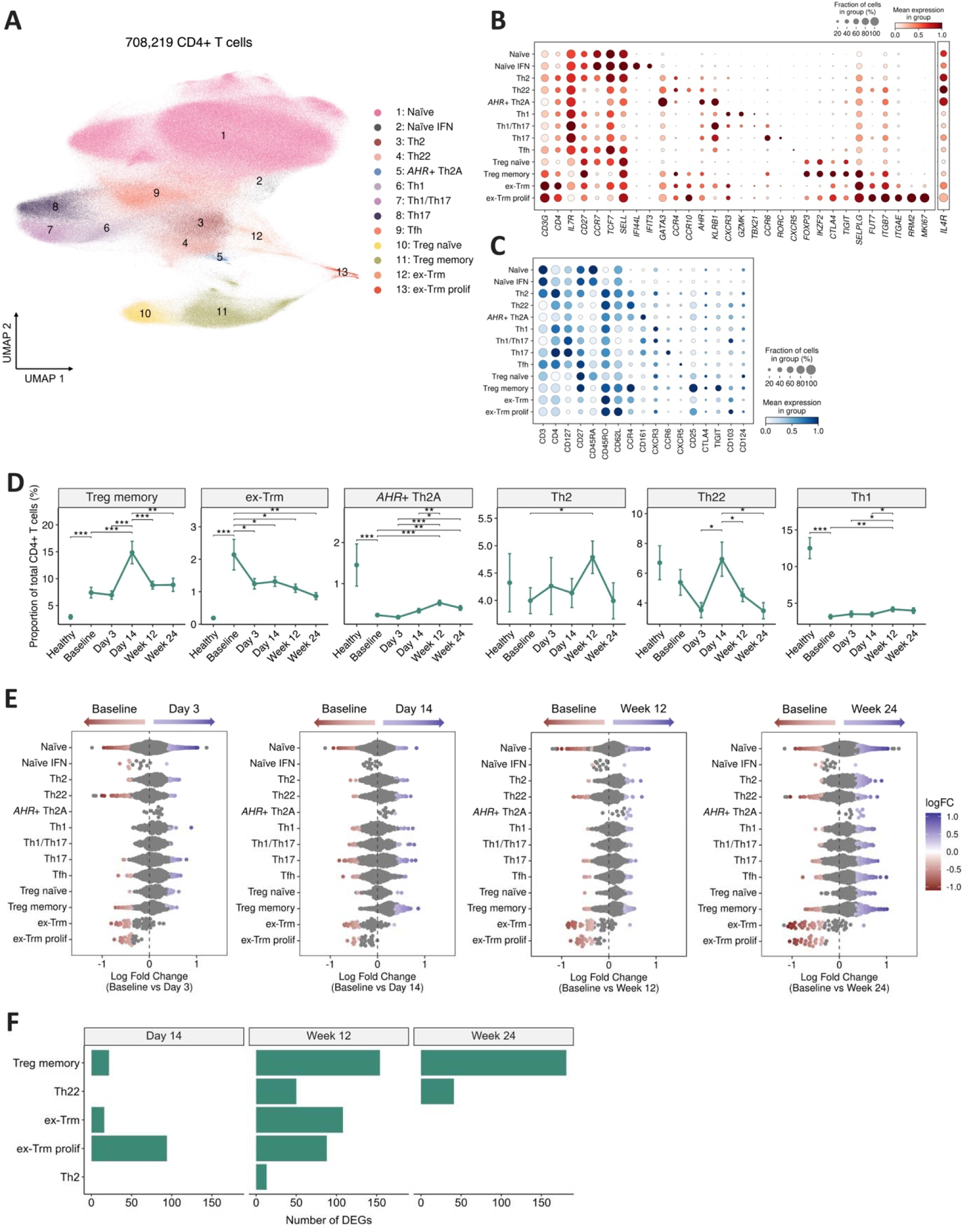
Changes in circulating CD4+ T cells during drug-induced AD resolution. **A)** UMAP visualization of 708,219 CD4+ T cells colored by cell type. **B-C)** Dot plots of CITE-seq RNA (B) and protein (C) readouts, showing the expression of the markers used to annotate CD4+ T cell subsets. **D)** Line plots of the memory T cell populations showing significant changes in abundance after treatment. Data are presented as mean +/- SEM. \**P*<0.05; \*\**P*<0.01; \*\*\**P*<0.001 (mixed design ANOVA for time-point analysis; Wilcoxon rank-sum test for comparison between healthy controls and baseline AD, see Methods for further details). **E)** Beeswarm plot showing the results of the Milo analysis. Cell neighborhoods with significant differences in abundance across time points (spatial FDR<0.1) are colored in shades of red (abundance increased at baseline) or blue (abundance increased after treatment). **F)** Bar plots showing the number of DEG observed at the indicated timepoints vs baseline. No DEG were detected on day 3.

### The circulating CD4 T-cell compartment undergoes significant changes during AD resolution

We compared the abundance of each circulating leukocyte population at day 0, day 3, day 14, week 12 and week 24 of treatment. We found no significant differences among CD8+ T cells (fig. S3d), B cells (fig. S4d), myeloid or plasmacytoid dendritic cells (fig S6d). We detected limited shifts in NK cells, where treatment caused a progressive increase in the abundance of a CD56^bright^ population (fig. S5d). Interestingly this subset has been characterized as a regulatory-like population that contributes to immune suppression ^21^.

At the same time, we observed that the frequency of various CD4+ T cell subsets changed during AD resolution, with the most prominent drug-induced shifts observed among memory Treg and CD4+ ex-Trm cells (Fig. 2d, fig. S3e). To validate these findings, we examined cell abundance using Milo, a statistical framework that analyses overlapping cell neighborhoods rather than discrete cell clusters ^22^. This confirmed that the resolution of inflammation was accompanied by significant changes in the proportion of memory Treg and CD4+ ex-Trm cells. The Milo analysis also uncovered early post-treatment shifts in the abundance of proliferating ex-Trm cells, Th22 cells and Th2 cells expressing *AHR* (Fig. 2e).

The latter population was positive for *KLRB1*/CD161, *PTGDR2* and *HPGDS* (Fig. 2b-c and Fig. S7a). This pattern matches the profile of Th2A cells, a proallergic Th2 subset that has been implicated in the pathogenesis of AD and asthma ^23^. However, the cells observed in our dataset also expressed *AHR* (Fig. S7a), a ligand-activated transcription factor that promotes skin barrier function and Treg differentiation ^24^. This suggests that the *AHR*+ Th2A population may have an immune regulatory role. Accordingly, we observed that the abundance of these cells was inversely correlated to eczema severity (r=-0.5; *P*=1.8x10^-6^) (Fig. S7b).

To complement the cell abundance analysis, we compared gene expression levels across time points, in all circulating CD4+ T cell subclusters. We found that expression changes were most conspicuous in the cell populations that also showed shifts in abundance (Fig. 2f). The *AHR*+ Th2A cells were the only exception to this pattern. We could not detect any differentially expressed genes (DEG) in this population, most likely owing to the small number of cells in the dataset (<3,000 cells across five timepoints).

Thus, our findings identify memory Treg, Th22, CD4+ ex-Trm and proliferating ex-Trm cells as the populations that are most affected during drug-induced AD resolution. We therefore decided to investigate these immune subsets in further detail.

### Drug-induced remission is associated with an increase in the abundance of memory Treg cells

The abundance of circulating memory Treg cells (identified as a CD45RA-/CD45RO+ Treg population, Fig. 1b-c) increased sharply after only two weeks of treatment (day 14), reducing to near-baseline levels as clinical remission ensued (week 12-24). Accordingly, the abundance of naïve Treg cells dropped on day 14 and rebounded in week 12 (Fig. 1d). The transient nature of these shifts was also confirmed with Milo (fig. S8a). Of note, clonal diversity was unchanged across timepoints (fig. S8b), suggesting that the increase in cell abundance was not driven by the expansion of a small number of clones.

Limited expression changes were observed on day 14 (Fig. 2f). Conversely, 154 DEG were detected at week 12. A marked downregulation of T cell activation and T cell migration pathways was observed (Fig. 3a-b), in line with the resolution of disease activity.

**Fig. 3.**
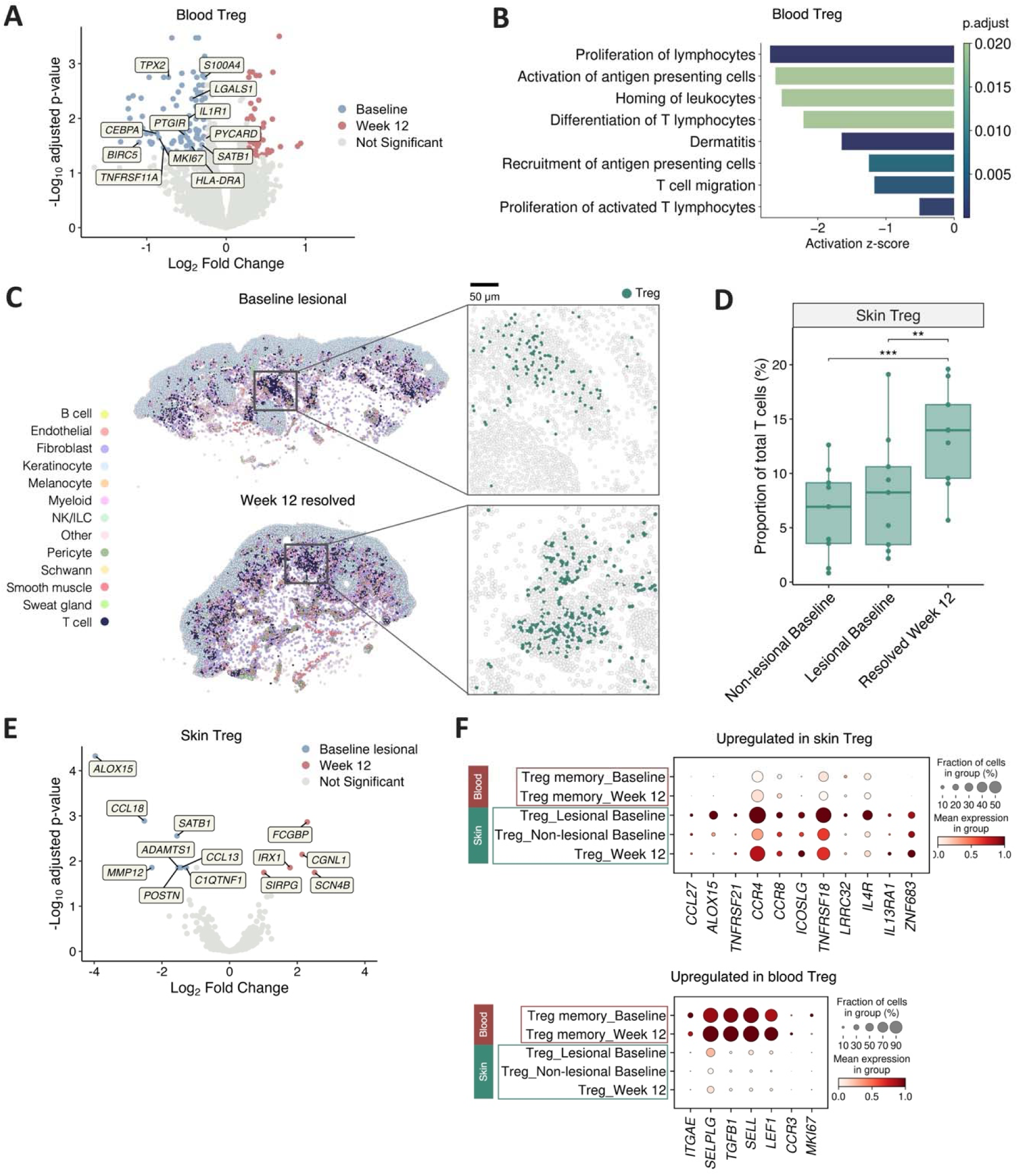
Treg cell changes induced by IL4Rα blockade during AD resolution. **A)** Volcano plot showing genes that are differentially expressed (log2(fold change)>|log2(1.2)|, *FDR*<0.05) in circulating memory Treg cells after 12 weeks of treatment. Colored dots represent genes that are upregulated at baseline (blue) or week 12 (red). Labels show selected inflammatory genes. **B)** Results of pathway enrichment analysis for the differentially expressed genes shown in A. **C)** Spatial transcriptomics of skin Treg cells. Left: representative images of skin sections where cells are colored based on their identity. Right: enlarged view showing Treg cells in dark green. **D)** Box plots showing treatment-induced changes in the abundance of skin Treg cells. \*\**P*<0.01; \**P*<0.001 (mixed design ANOVA, followed by pairwise comparison of estimated marginal means and Bonferroni correction). **E)** Volcano plot showing genes that are differentially expressed in skin Treg cells at week 12 vs baseline. Genes yielding a log2(fold change)>|log2(1.2)| and an *FDR*<0.05 are shown by colored dots with name labels. **F)** RNA expression dot plots showing significant differences between skin and circulating Treg cells.

To follow up these findings in skin, we analyzed spatial transcriptomic data from nine study participants, as reported elsewhere ^15,25^ (fig. S8c-d). By comparing cell frequencies at baseline and week 12 of treatment, we found that the abundance of skin-resident memory Treg cells also increased with the resolution of inflammation (Fig. 3c-d). The expansion was accompanied by the downregulation of inflammatory molecules (*CCL13* and *CCL18*) and the reduced expression of *SATB1*, a transcription factor that antagonizes Treg cell differentiation ^26^ (Fig. 3e). These observations mirrored changes in blood, i.e. the downregulation of *SATB1* and the modulation of inflammatory pathways, observed in circulating Treg cells after 12 weeks of treatment (Fig 3a-b).

To further explore these findings, we compared Treg expression profiles in blood and skin. In keeping with published studies ^27^, we found substantial similarities between the Treg genes expressed in the two compartments (fig. S8d). However, we also observed some important differences. Compared to their circulating counterparts, skin-resident Treg cells upregulated genes that are important for their immune suppressive activity (*ALOX15* ^28^), tissue residency (*ZNF683*, encoding the transcription factor hobit ^29^) and the response to chemoattractants produced by migratory DCs (*CCR4* ^30^) (Fig 3f).

Taken together, these analyses show that IL4Rα blockade induces an expansion of circulating and skin-resident memory Treg cells. In blood, this is an early, transient phenomenon that precedes clinical remission. Conversely, the increase in skin Treg abundance is sustained for at least 12 weeks, reflecting an expression profile that supports Treg retention and stability during disease resolution.

### The abundance of CD4+ ex-Trm cells rapidly decreases during drug-induced remission

Following IL4Rα blockade, the resolution of inflammation was accompanied by a rapid (day 3) and significant reduction in the abundance of circulating CD4+ ex-Trm cells (Fig. 2d-e). These represent a rare population (1-2% of CD4+T cells in our dataset), characterized by the robust expression of skin homing receptors (e.g. *CCR4*, *CCR10* and *SELPLG,* which encodes CLA) and skin residency markers (*ITGAE*/CD103, *ITGB7*) (Fig. 4a). A similar profile was observed in a second ex-Trm population, which displayed a strong upregulation of cell proliferation markers (*RRM2* and *MKI67)* (Fig. 4a).

**Fig. 4.**
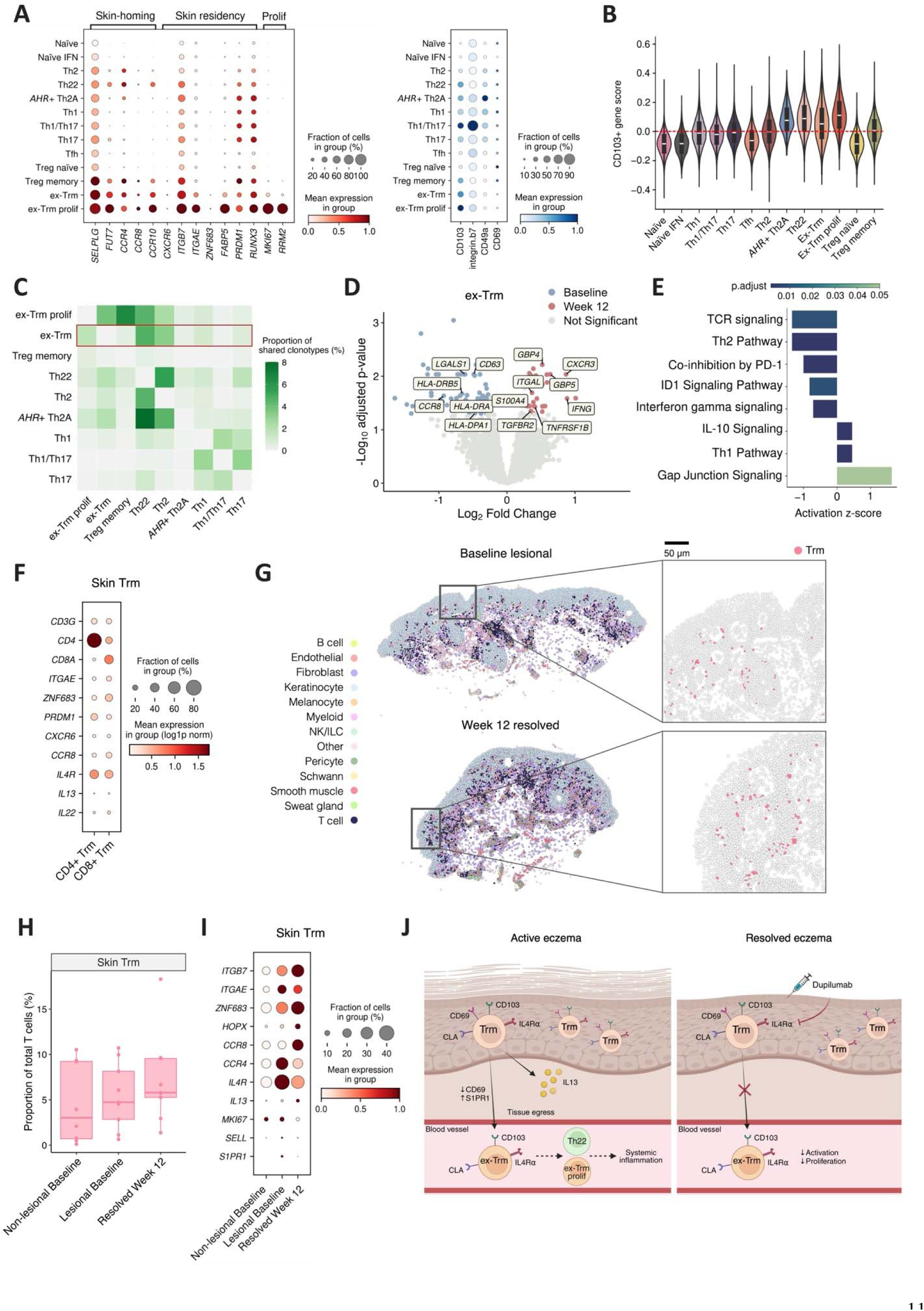
Changes in circulating CD4+ ex-Trm cells and skin Trm cells during AD remission. **A)** Dot plots of PBMC CITE-seq showing RNA (left) and protein (right) expression of markers used to annotate CD4+ ex-Trm cells. **B)** Violin plots showing the aggregated expression of genes that are differentially expressed in CD103+ T cells (CD103 gene score). **C)** Heatmap showing the percentage of shared TCR clonotypes between pairs of circulating CD4+ T cell populations. To improve clarity, T cell subsets sharing <2% of their clonotypes with other populations were omitted. **D)** Volcano plot showing genes that are differentially expressed (log2(fold change)>|log2(1.2)|, *FDR* <0.05) in circulating CD4+ ex-Trm cells after 12 weeks of treatment. Colored dots represent genes that are upregulated at baseline (blue) or week 12 (red). Labels show selected inflammatory genes. **E)** Results of pathway enrichment analysis for the differentially expressed genes shown in D. **F)** Dot plot showing the RNA expression of genes used to annotate skin Trm cells. **G)** Spatial transcriptomics of skin Trm cells. Left: images of skin sections where cells are colored based on their identity. Right: high resolution inset with cells colored as above (left panel) or Trm cells shown in pink (right panel). **H)** Box plots showing the abundance of skin Trm cells at different time points. **I)** Dot plot showing longitudinal RNA expression changes detected in skin CD4+ Trm cells. **J)** Schematic showing the proposed dynamics of CD4+ ex-Trm cells during AD resolution. The dotted arrow denotes a relationship inferred from shared cell clonality (image created with biorender.com).

Interestingly, the expression of the CD103 skin residency marker was also detectable in Th22 and *AHR+* Th2A cells (Fig. 4a). To further investigate this observation, we measured an ex-Trm transcriptional signature, defined by Klicznik et al through the characterization of sorted CD4^+^CLA^+^CD103^+^ T cells ^3^. We found that the signature was prominently expressed in the two ex-Trm populations, but also in Th22 and *AHR*+ Th2A cells (Fig. 4b).

Given the phenotypic plasticity of ex-Trm cells ^5^, this suggested that an original recirculating population may have differentiated along multiple axes, acquiring a proliferative, Th22 or *AHR*+ Th2A phenotype. This hypothesis was supported by the observation of TCR clonotype sharing between CD4+ ex-Trm, proliferating ex-Trm, Th22 and *AHR+* Th2A cells (Fig. 4c).

We next compared gene expression at week 12 of IL4Rα blockade vs baseline. While we did not observe any DEG in the small *AHR+* Th2A population, our analysis showed that treatment had comparable effects on ex-Trm, proliferating ex-Trm and Th22 cells. In these subsets, the resolution of inflammation was accompanied by the downregulation of genes related to T cell activation (e.g. *HLA-DRA* and *HLA-DPA1*) and skin homing (e.g. *CCR8* or *CCR10*) (Fig 4d and Fig S9a), leading to reduced TCR signaling and Th2 activation (Fig. 4e).

To further investigate these findings, we examined drug-induced changes in skin Trm cells. The analysis of patient biopsies showed that the Trm population consisted almost exclusively of *IL13+* cells (fig. S9b), which expressed skin residency (*ITGAE*, *ZNF683*) and skin homing (*CCR8*) markers (Fig. 4f). In keeping with reports of Trm cell persistency in resolved AD skin ^31^, we found that the abundance of these cells had not significantly changed by week 12 of treatment, when clinical remission was apparent (Fig. 4g-h). We next investigated skin CD4+ Trm gene expression during the resolution of inflammation (week 12 vs baseline). We found that genes mediating immune suppression (*HOPX, TIGIT*^32^) and skin residency (*ZNF683*)^29^ were upregulated during IL4Rα inhibitor treatment (Fig. 4i).

These findings show that a small fraction of CD4+ Trm cells leave inflamed skin to recirculate in the bloodstream. Our TCR-seq analysis further suggests that ex-Trm cells may differentiate into T helper subsets that have been implicated in AD pathogenesis (e.g. Th22 cells ^7^). Of note, the egress of Trm cells from skin is reduced during drug-induced AD resolution. As IL4Rα blockade dampens inflammation, the expression of Trm genes mediating skin residency is reinforced and the abundance of circulating ex-Trm cells decreases, well in advance of clinical remission (Fig. 4j).

### The abundance of circulating CD4+ ex-Trm cells correlates with disease activity

To further investigate the link between disease activity and the frequency of CD4+ ex-Trm cells, we took three different approaches. First, we examined our PBMC CITE-seq dataset. This showed that the abundance of CD4+ ex-Trm and proliferating ex-Trm populations strongly correlated with Eczema Area and Severity Index (EASI), the gold standard measure of AD disease activity (r ≥0.35; *P* ≤0.0015) (Fig. 5a). Next, we used flow cytometry to analyze an independently ascertained cohort including nine individuals with moderate-to-severe AD, who had been successfully treated with dupilumab (fig. S1b, table S2). After gating CD4+ ex-Trm cells as CD4+CLA+CD103+ T cells (fig. S10a), we were able to confirm that the abundance of this population decreased early (day 14) during the resolution of skin inflammation, well in advance of clinical remission (Fig. 5b-c). Finally, we sought to demonstrate that these changes are reversed during the recurrence of inflammation. Thus, we recruited eight AD patients who had discontinued dupilumab treatment after achieving durable remission (clear skin for >1 year) (table S3). We examined blood samples collected on the day of the first missed dose (baseline) and at regular intervals in the run up to clinical recurrence (Fig. 5d; fig. S10b). The analysis of single-cell transcriptomes and protein surface expression showed that the frequency of CD4+ ex-Trm cells consistently increased with the return of first skin symptoms (Fig. 5e). Thus, changes in the abundance of CD4+ ex-Trm cells closely mirror the evolution of skin inflammation during AD remission and recurrence.

**Fig. 5.**
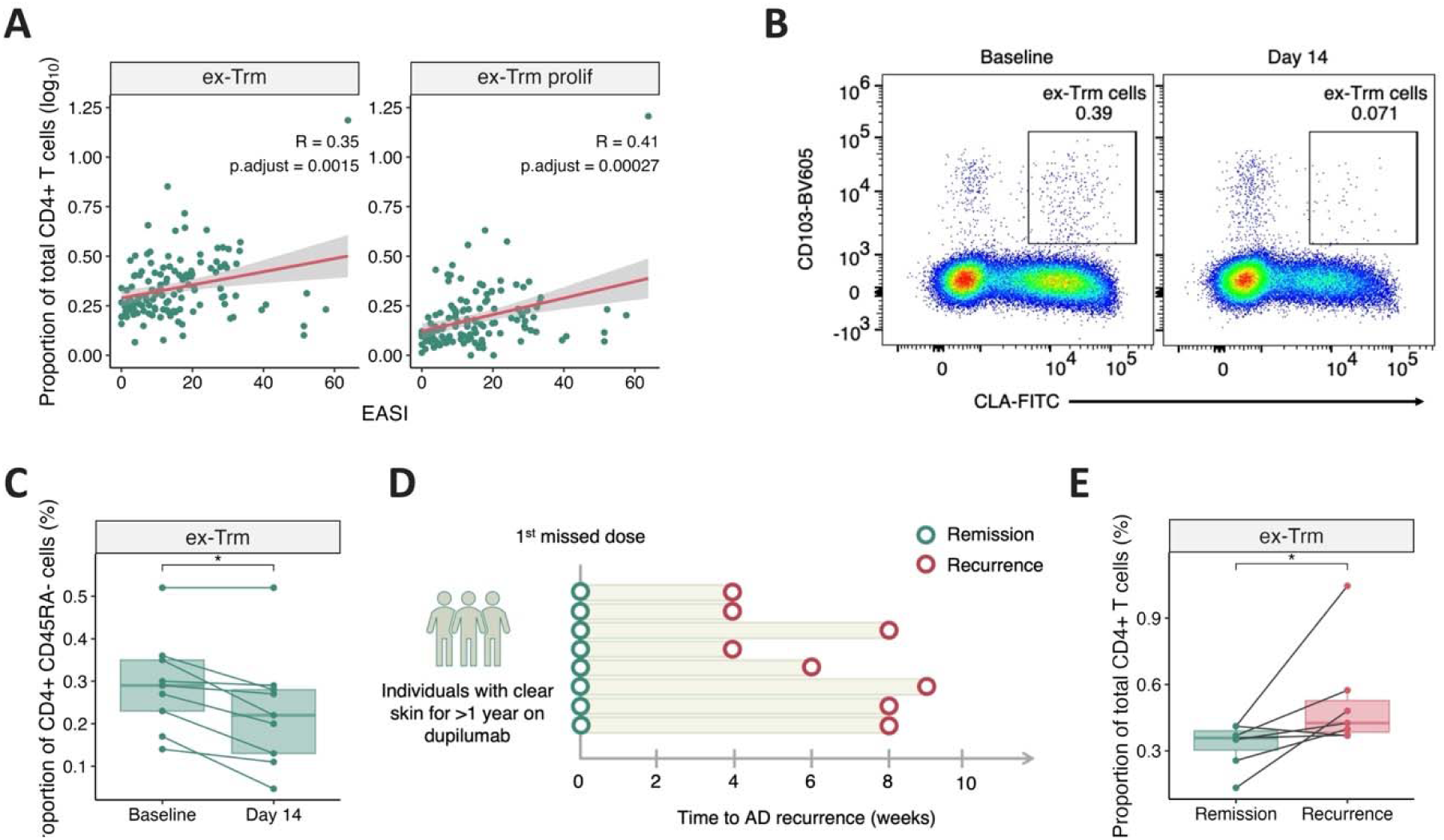
Correlation between the abundance of circulating CD4+ ex-Trm cells and AD disease activity. **A)** Correlation between disease activity and the abundance of ex-Trm populations. Each dot is a sample; the regression line is shown in red with 95% confidence intervals shaded in grey. The p-value was calculated by repeated measure correlation analysis and adjusted using the Benjamini-Hochberg correction. EASI, eczema area and severity index. **B-C)** Flow cytometry analysis of CD4+ ex-Trm cells. B) representative flow cytometry plot showing reduced CD4+ ex-Trm abundance at day 14 of treatment, compared to baseline. C) Line and box plots showing longitudinal changes in CD4+ ex-Trm cell abundance measured in nine AD patients treated with dupilumab. Each line represents an affected individual. \**P*<0.05 (Wilcoxon signed rank test). **D)** Overview of the recurrence cohort, showing the time points when symptoms started to return. **E)** Line and box plots showing the abundance of CD4+ ex-Trm cells on the day of the first missed dose (two weeks after the last day of treatment, remission) and at the time of recurrence. \**P*<0.05 (Wilcoxon signed rank test).

## DISCUSSION

The aim of our study was to characterize the immune changes that occur during AD remission and recurrence. To this end, we generated a uniquely powered single-cell dataset, including ∼1.3M PBMCs sampled following commencement and withdrawal of IL4Rα blockade. This resource was complemented by single cell profiles of healthy donors and spatial transcriptomic data from longitudinal AD skin biopsies.

Our analysis of these datasets uncovered two critical immune shifts. First, drug-induced AD resolution is associated with the expansion of circulating and skin-resident immune regulatory populations. Second, clinical remission is preceded by early changes in circulating CD4+ ex-Trm cells, which are rapidly reversed upon drug-withdrawal and disease recurrence.

The role of immune regulatory cells during remission was foreshadowed by the changes we detected in an immune modulatory NK cell subset. In keeping with published findings, we found that total NK cell abundance was reduced in affected individuals compared to healthy donors ^17,18^. While others have shown that IL4Rα blockade restores NK cell numbers ^17,18^, our single-cell analysis demonstrated that treatment specifically upregulates a CD56^bright^ NK subset, characterized by high expression of granzyme K. These cells contribute to immune tolerance by producing anti-inflammatory cytokines and targeting activated CD4+ T lymphocytes ^21^. They are mostly found in peripheral organs, but their tissue residency is transient, as they continuously recirculate in the blood ^33^. Thus, the shift from inflammation to remission could affect CD56^bright^ NK cell dynamics, altering the balance between extravasation, skin residency and egress.

Our analysis of CD4+ T cell subclusters also showed that clinical remission was preceded by a transient increase in the abundance of circulating memory Treg cells. This is consistent with the results of a flow cytometry study, carried by Bakker et al, in a small longitudinal dataset ^9^.

Interestingly, agents that promote Treg expansion are being investigated as potential AD therapeutics ^34,35^ and promising results have been reported in a clinical trial of rezpegaldesleukin, a Treg-selective IL-2 receptor agonist ^35^. In this context, we show that the early and transient expansion of circulating Tregs induced by IL4Rα inhibition is accompanied by a progressive increase in the abundance of their skin-resident counterparts. Following treatment, these cells downregulate Th2-related markers (*CCL13, CCL18* and *ALOX15*), thus modulating the Th2-like phenotype that characterizes AD Tregs ^36^. The expansion of skin Tregs may then be sustained by their proximity to migratory DCs, a specialized population that promotes Treg generation within draining lymph nodes ^15,37^.

Our analysis of CD4+ T cell subclusters also identified a population of Th2 cells that was characterized by strong expression of *AHR,* the gene encoding the aryl hydrocarbon receptor. In affected individuals, the abundance of *AHR*+ Th2A cells was reduced compared to healthy controls and inversely correlated with AD disease activity. These observations are in keeping with the immune modulatory role of AHR signaling, which is now being harnessed through the therapeutic use of receptor agonists in AD ^24^. Thus, immune modulatory treatment restores a variety of anti-inflammatory pathways, mediated by NK cells, and by memory subsets such as Tregs and *AHR*+ Th2A cells.

The second, key observation emerging from our study is that changes in the egress of Trm cells from the skin are early indicators of clinical remission and recurrence.

As expected ^31^, we found that treatment did not affect the abundance of skin Trm cells. Conversely, these cells showed an increase in the expression of *ZNF683*/Hobit (a master regulator of lymphocyte tissue residency), during drug-induced AD resolution ^29^. While previous studies have suggested that inflammation downregulates genes that mediate residency ^5^, our findings indicate that this phenomenon can be reversed by treatment that restores skin immune homeostasis. Of note, Strobl et al have shown that the mobilization of skin ex-Trm cells can propagate inflammation to distant sites ^38^. This is in keeping with our TCR-seq results, indicating that CD4+ ex-Trm cells may differentiate into Th22 cells, a population that sustains chronic skin inflammation ^7^. In this context, it is tempting to speculate that early intervention in AD might reduce the pathogenic potential of Trm egress from skin, with beneficial effects on the overall disease trajectory.

Ramirez-Navarro et al recently described a population of CD8+ ex-Trm cells, recirculating from the gut of healthy donors and individuals with Crohn’s disease. These cells expressed residency markers (*ITGAE*/CD103, *ITGA1*) and gut-, rather than skin-homing receptors (*CCR9*) ^4^. Interestingly, our unbiased clustering identified a similar subset of *CCR9+*/CD103+ CD8+ T cells in our dataset, providing independent confirmation for the existence of this population.

Ramirez-Navarro et al also showed that Crohn’s disease endoscopic scores correlated with expression signatures in CD8+ ex-Trm cells, indicating that changes in these cells mirror disease activity in their tissue of origin ^4^. We obtained similar results, showing that the abundance of CD4+ ex-Trm cells correlated with clinical AD activity (EASI). We further demonstrated that the proportion of CD4+ ex-Trm cells starts to drop after only three days of treatment – well in advance of clinical remission. In individuals who previously achieved durable remission, this change is reversed following drug withdrawal, with cell abundance rebounding upon the return of the first skin symptoms.

Our study has some limitations. We only carried out TCR-sequencing in PBMCs, so we could not track the migration of T cell clones between skin and blood. Since IL4Rα blockade is highly effective, our prospectively recruited cohorts only included individuals who had achieved a clinical response following treatment. While the focus of this study was on drug-induced remission, we acknowledge that the analysis of non-responders could have provided further validation for our results (e.g., by showing that there is no reduction in CD4+ ex-Trm abundance, in individuals who do not respond to treatment).

In conclusion, we have shown that early changes in the abundance of circulating CD4+ ex-Trm cells precede clinical remission and recurrence in AD. While these results will require validation in further datasets (including cohorts treated with other immune modulators), they suggest that CD4+ ex-Trm/Trm cell dynamics play a central role in AD remission and recurrence. In the future, these cells could be investigated as therapeutic targets for disease modification and biomarkers of drug response, with potential to inform personalized treatment strategies guided by immune monitoring.

## METHODS

### Patient recruitment and sample collection

This study was undertaken in accordance with the principles of the Helsinki declaration. Written informed consent was obtained from all participants, in line with approval from the London – Westminster Research Ethics Committee (REC ref EC00/128). Affected individuals were recruited via the national specialized service for AD at St John’s Institute of Dermatology (Guy’s and St Thomas’ NHS Foundation Trust) or via the BEACON clinical trial (ISRCTN11056540). The main study cohort (table S1) included 35 adults who had been diagnosed with moderate-to-severe AD by a dermatologist. Most (34/35; 97%) were biologic naïve and all received dupilumab as a monotherapy, achieving a 50% EASI reduction by week 24. Blood was collected from all 35 participants at baseline (day 0), day 14 and week 12 of treatment, with additional samples taken on day 3 and week 24 in 23 individuals. Full-thickness skin biopsies were obtained from 9 participants at baseline (lesional and non-lesional skin) and week 12 (resolved lesional skin) ^15,25^. The flow cytometry cohort (table S2) included nine additional adult patients who were receiving dupilumab as a monotherapy, as above. All were biologic naïve and donated blood samples on day 0 and day 14. The drug withdrawal cohort (table S3) included eight adults with moderate-to-severe AD, who had achieved durable remission (clear or nearly clear skin (EASI ≤ 1.5) for >1 year) and discontinued dupilumab as part of routine clinical care. Blood samples were collected two weeks after the last day of treatment (i.e. on the day of first missed dose) and at regular intervals until recurrence (defined as the earliest indication that an area of eczema had returned). In these individuals, dupilumab treatment was restarted immediately following recurrence.

### Sample processing

PBMCs were isolated by gradient centrifugation and stained with the TotalSeq-C antibody cocktail (BioLegend, 99813) according to the manufacturer’s instructions. Cells were processed using the Chromium Next GEM Single Cell 5’ Reagent Kit v2 (Dual index) with Feature Barcoding technology for Cell-Surface Protein and Immune Receptor Mapping (Rev A protocol; 10x Genomics). Cells were loaded on a Chromium chip (10x Genomics) and gene expression, cell-surface protein, and TCR-enriched libraries were pooled at an 8:1:1 ratio. Libraries were then sequenced with Illumina Novaseq 6000 or Illumina Novaseq X instruments.

Skin biopsies were immediately frozen in OCT. Tissue sections (10 µm) were placed on a Xenium slide (10x Genomics) and in situ gene expression data was generated using the Xenium in situ 5k-plex platform (10x Genomics), according to the manufacturer’s protocol.

### CITE-seq data processing

FASTQ files were processed using Cellranger multi (v7.1.0) (10X Genomics) and reads were aligned to the GRCh-2020-A reference genome. Ambient mRNA reads were removed with CellBender (v.0.03) ^39^ and Scrublet (v0.2.3) ^40^ was applied to the RNA modality of each sample separately to generate a doublet score per cell. Cells where the doublet score exceeded the sample median by >3 median absolute deviations were removed. All subsequent steps were performed with Scanpy (v1.11.4) ^41^. For quality control, cells with <200 or >6000 UMI counts, and cells with >6% mitochondrial reads were removed. Genes detected in <3 cells were also eliminated. Finally, cells were removed if the protein library size or number of detected proteins exceeded the sample median by more than 3 median absolute deviations. Total counts for the remaining cells were normalized to 10,000 and log transformed. Highly variable genes (HVGs) were identified using the *highly_variable_genes* function in Scanpy with the Seurat v3 algorithm (minimum mean variance = 0.0125, maximum mean variance = 3, minimum dispersion = 0.5). Surface protein and RNA counts were integrated using totalVI (scvi-tools package, v1.3.3) ^42^, using a model trained on the top 5000 HVGs with a 20-dimensional latent space, 2 decoder layers and a learning rate of 4x10^-3^ for 300 epochs. A joint latent representation with denoised protein and RNA values was generated with sequencing run as batch key and donor as a technical covariate. A reinspection of clinical records showed that one study participant had very low baseline disease activity (EASI <1.5). The cells of this individual were kept in the dataset to facilitate clustering and sub-clustering of immune populations. They were, however, excluded from all differential abundance and differential expression analyses.

### Cell clustering and annotation

A k-nearest neighbor graph (k=30) was constructed in Scanpy, based on totalVI RNA and protein embeddings. Initial cell clustering was implemented using the Leiden algorithm at a resolution of 1.6. For manual annotation of broad cell identities, the DEG defining each cluster (identified with the *sc.tl.rank_ genes_groups* function in Scanpy) were compared with human PBMC markers from the Azimuth reference dataset ^43^. Annotations were then validated with CellTypist (v1.7.1) ^44^, using the majority voting approach to interrogate two models, each trained on a healthy PBMC dataset ^16,45^.

Broad cell types were separated into distinct CD4+ T cell, CD8+ T cell, B cell, NK cell and monocyte objects. Post-filtering raw counts for each object were normalized and log transformed. Joint RNA and protein embeddings were generated using totalVI, with sequencing run as batch key and donor as a technical covariate. Nearest neighbor graphs (k=20) were constructed and sub-clusters defined using the Leiden algorithm with 0.8-2.0 resolution values. Cell identities were manually annotated by cross-referencing DEG (identified as above) with markers described in the literature. Annotations were validated using decoupleR (v2.0.6) ^46^ to score cells against CellMarker 2.0 Human Normal Blood reference gene sets ^47^.

### Integration of publicly available datasets

scRNA-seq data generated in PBMCs ^16^ was downloaded from ArrayExpress (study id: E-MTAB-10026). Healthy control data were extracted and processed using the QC filters described above. The data were first integrated with our dupilumab dataset using scVI ^48^ (scvi-tools package) with the top 2000 HVGs, with ‘Dataset’ as the batch key and donor as a covariate. The trained scVI model was then used to initialize a scANVI model ^49^ (scvi-tools package) to perform semi-supervised cell type annotation in healthy controls, using the annotation of the dupilumab dataset as reference (fig. S11).

### Differential abundance and differential expression analysis

Compositional analysis of the entire blood compartment was implemented with scCODA (v0.1.9) ^50^, using CD4+ naïve cells as the reference population. This framework enabled us to address the negative correlation between cell type proportions (i.e. the fact that cell type fractions are not independent from each other as they sum up to 1).

Neighborhood-based differential abundance analysis was performed using MiloR (v2.2.0) ^22^. A generalized linear mixed model, with donor as a random effect, was used to analyze abundance at the indicated timepoints. Significant neighborhoods were defined as those with a spatial *FDR*<0.1.

For differential expression analysis, pseudo-bulking was performed by aggregating raw gene counts across all cells from the same patient, cell type and timepoint. After filtering out samples with <10 cells, genes with <3 counts per sample, mitochondrial and ribosomal genes, generalized linear mixed models for repeated measures were constructed, using Dream (variancePartition package, v1.32.5) ^51^. To minimize confounders, the following co-variates were included: sex, age, total number of aggregated cells, percentage of mitochondrial genes, sequencing batch and donor. Gene set enrichment analysis was performed with Ingenuity Pathway Analysis package (Qiagen). Cells were scored for CD103 expression signatures using the *sc.tl.score_genes* function, based on a set of genes that were up- or downregulated in CD4+CLA+CD103+ cells ^3^.

### TCR-seq data processing and analysis

The Cell Ranger VDJ pipeline (10x Genomics) was used to align and assemble T-cell Receptors (TCRs). Indexing and quality control was performed using Scirpy (v.0.16.1) ^52^. Cells with >2 receptor pairs (multichain) or with a single receptor sequence (orphan chain) were excluded from analysis. The remaining productive TCRs and cell barcodes were matched to post-filtering CITE-seq data. TCR sharing was investigated by identifying clonotypes that originated from different cell types but exhibited identical CDR3 sequences and V/J gene usage. Repertoire similarity was first assessed using the *scirpy.tl.repertoire_overlap* function. For each cell type, the number of clonotypes that were also present in other cell populations (shared clonotypes) was quantified as a percentage of the total number of clonotypes.

### Xenium data processing and analysis

Spatial transcriptomic analysis was undertaken as previously described ^53^.

The Xenium data was integrated with the CITE-seq profiles (RNA modality) of the relevant patients. scVI was run with the top 2000 highly variable genes, ‘Tissue’ as the batch key and donor as a covariate. A k-nearest neighbor graph was constructed (k=30) and clustering was performed using the Leiden algorithm with resolution values ranging from 0.8 to 2.0 in increments of 0.2. Partition-based graph abstraction (PAGA) was computed on the KNN graphs with the *sc.tl.paga* function in Scanpy and used for UMAP calculation.

Differences in gene expression between skin and blood Tregs were analyzed using edgeR (v4.0.16) ^54^. Pseudo-bulk gene expression was derived by aggregating cells by patient and timepoint. Samples with less than 10 cells and genes expressed at low levels (min.count = 3, min.total.count=5) were removed and read counts were normalized using trimmed mean of M-values (TMM). Differential expression testing was performed using a quasi-likelihood negative binomial model with donor as a covariate. P-values were adjusted using the Benjamini-Hochberg correction and genes achieving log2(fold change)>|l.0| with an *FDR*<0.05 were considered differentially expressed.

### Flow cytometry

PBMCs were processed using the CD4+ T cell isolation kit (Miltenyi Biotec) according to the manufacturer’s instructions. CD4+ T cells were then incubated with LIVE/DEAD™ Fixable Aqua stain (Invitrogen) for 30 minutes at 4°C and with Human TruStain FcX Fc receptor blocking solution (BioLegend) for 10 minutes at room temperature. Cells were stained with the antibodies of interest (table S4) in FACS buffer (PBS with 2% FBS and 2mM EDTA) and Brilliant Stain Buffer (ThermoFisher Scientific) (1:1 v/v), for 30 minutes at 4°C. Cells were acquired on a CytoFLEX flow cytometer (Beckman Coulter). Data was analyzed using FlowJo (BD, v10.10), after performing quality control with PeacoQC ^55^.

### Statistical analysis

The correlation between cell abundance and EASI scores was calculated using rmcorr (v0.7.0) to account for repeated measures ^56^. P-values were then adjusted using the Benjamini-Hochberg correction. Comparison of cell abundance between different time points was performed using a mixed design ANOVA with donor as a random effect. A pairwise comparison of estimated marginal means was implemented as a post-hoc test, with Benjamini-Hochberg adjustment of p-values. Comparison of CD4+ T cell abundance between healthy controls and baseline AD was performed using a Wilcoxon rank-sum test.

## Data availability

Full access to skin spatial data, PBMC CITE-seq and TCR-seq data will be provided by the time of publication via an online web portal.

## Acknowledgements

This research was supported by Research and Development, Guy’s and St Thomas’ NHS Foundation Trust. We are grateful for the support of St John’s Institute of Dermatology Skin Therapy Research Unit (John Gregory, Bindi Gaglani, Amelia Mitchell-Gears, Ilona Blee, Kingsley Powell, Jade Pizzato, Katherine Teather, Varshaa Anantharam, Ineta Andrijauskaite, April Qin Neville, Ifeoluwa Olabiyi, Majura Srikanthan, Veronika Krasna, Michael Duckworth, Tejus Dasandi, Maria Troy, Selina Cox, Sam Curtis, Andrew Lovell).

This research was also supported through funding from the King’s Health Partners Centre for Translational Medicine. The views expressed are those of the authors and not necessarily those of King’s Health Partners.

CHS is a National Institute of Health and Care Research (NIHR) senior investigator. The views expressed are those of the author(s) and not necessarily those of the NHS, the NIHR or the Department of Health and Social Care.

## Funding

This work was funded by grants from the National Institute of Health Research (NIHR151210 to SKM, CHS, FC and MH; NIHR129926 to CHS and AEP), Open Targets (OTAR3085 to MH, SKM, CHS), Leo Foundation (LF-OC-22-000935 to FC, SKM) and Wellcome Trust (316726/Z/24/Z to MS). TJ is supported by Medical Research Council (MRC) grant MR/W006820/1 (industry partner: Astra Zeneca) and King’s College London as a member of the MRC Doctoral Training Partnership in Biomedical Sciences. KD is supported by the Psoriasis Association (grant ST1/25) and JYWL by the Medical Research Council (grant UKRI2437). FC is supported by the BC Leading Edge Endowment Fund and BC Children’s Hospital Research Institute. SKM is funded by grant NIHR302258.

## Author contributions

M.H., C.H.S., F.C. and S.K.M. conceived the study, obtained the necessary funding and supervised patient recruitment, sample and data processing. R.W., A.E.P., C.H.S. and S.K.M. facilitated patient recruitment, acquired samples, and collected clinical metadata, with help from R.M. and J.Y.W.L.; D.B. isolated PBMCs and processed skin biopsies. A.R.F., E.S., K.E., T.B., V.R., B.R. and K.S. generated the CITE-seq and spatial transcriptomic datasets. T.J., L.S., M.S., A.V.P., L.W., A.B. and H.J. analyzed the data with input from V.B.M.S; K.D. carried out the flow cytometry experiments with help from T.J and input from A.R. and P.M; M.K.L. helped with data interpretation. F.C. and S.K.M. drafted the manuscript, which was then reviewed by T.J. and A.R.F. with input from all co-authors.

## Competing interests

FC has received grants and consultancy fees from Boehringer Ingelheim. SKM has received funding from AstraZeneca, and reports departmental income from AbbVie, Almirall, Eli Lilly, Leo, Novartis, Sanofi and UCB outside the submitted work. CHS reports departmental research funding from AstraZeneca, Boehringer Ingelheim, UCB and Sanofi outside the submitted work, and serves as an investigator in EU-IMI consortia involving multiple industry partners (see biomap-imi.eu and hippocrates-imi.eu for details). RW has been a consultant, advisory board member, investigator, and/or speaker and received travel support for participation in congresses and/or (speaker) honoraria and/or research grants from AbbVie, Almirall, Amgen, Boehringer Ingelheim, Bristol Myers Squibb, Celgene, Eli Lilly, Galderma, Incyte, Janssen-Cilag, LEO Pharma A/S, Novartis, Pfizer, Sanofi, and UCB. All other authors declare no competing interests.

## Notes

### Author Declarations

London - Westminster Research Ethics Committee gave ethical approval for this work (REC ref EC00/128).

