## Supplementary Figures and Tables for "Atopic dermatitis remission and recurrence are preceded by changes in ex-tissue-resident memory T cell abundance"

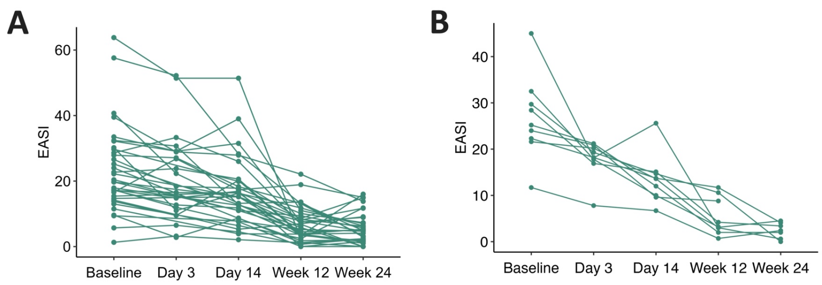


**Fig. S1. AD severity scores measured in study participants receiving IL4Rα inhibitory therapy. A-B)** Line plots showing longitudinal changes in Eczema Area and Severity Index (EASI) in AD patients analysed by CITE-seq and TCR-seq (n=35) (A) or flow cytometry (n=9) (B). Each line represents an affected individual.

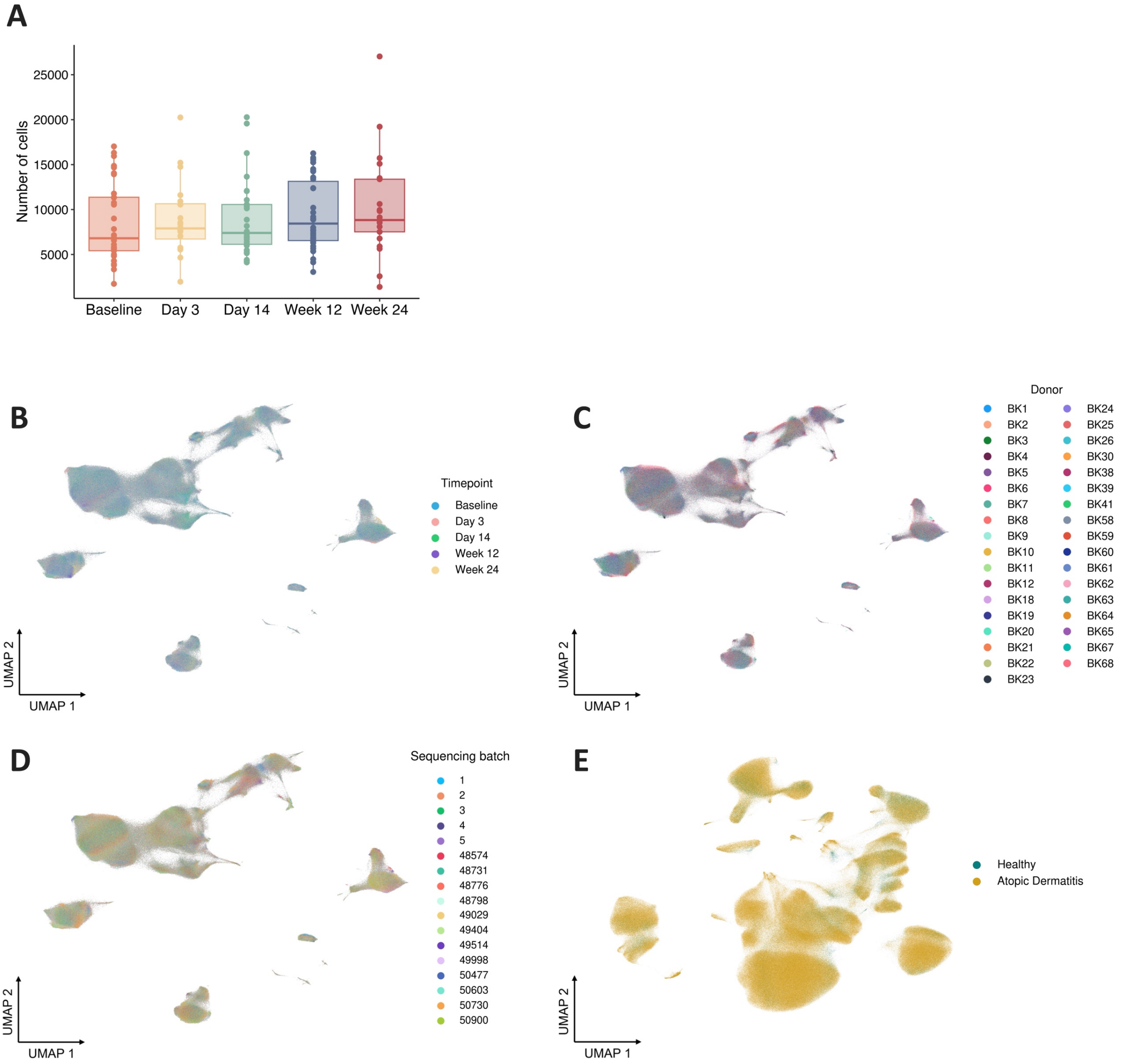


**Fig. S2. Quality control of data generated by Cellular Indexing of Transcriptomes and Epitopes by Sequencing (CITE-seq) A)** Box plots showing the number of cells that passed QC at each timepoint. Each dot represents a sample. **B-D)** UMAPs showing that cells do not cluster by timepoint (B), donor (C) or sequencing batch (D). **E)** UMAP showing the integration of the study cohort with a publicly available healthy PBMC dataset.

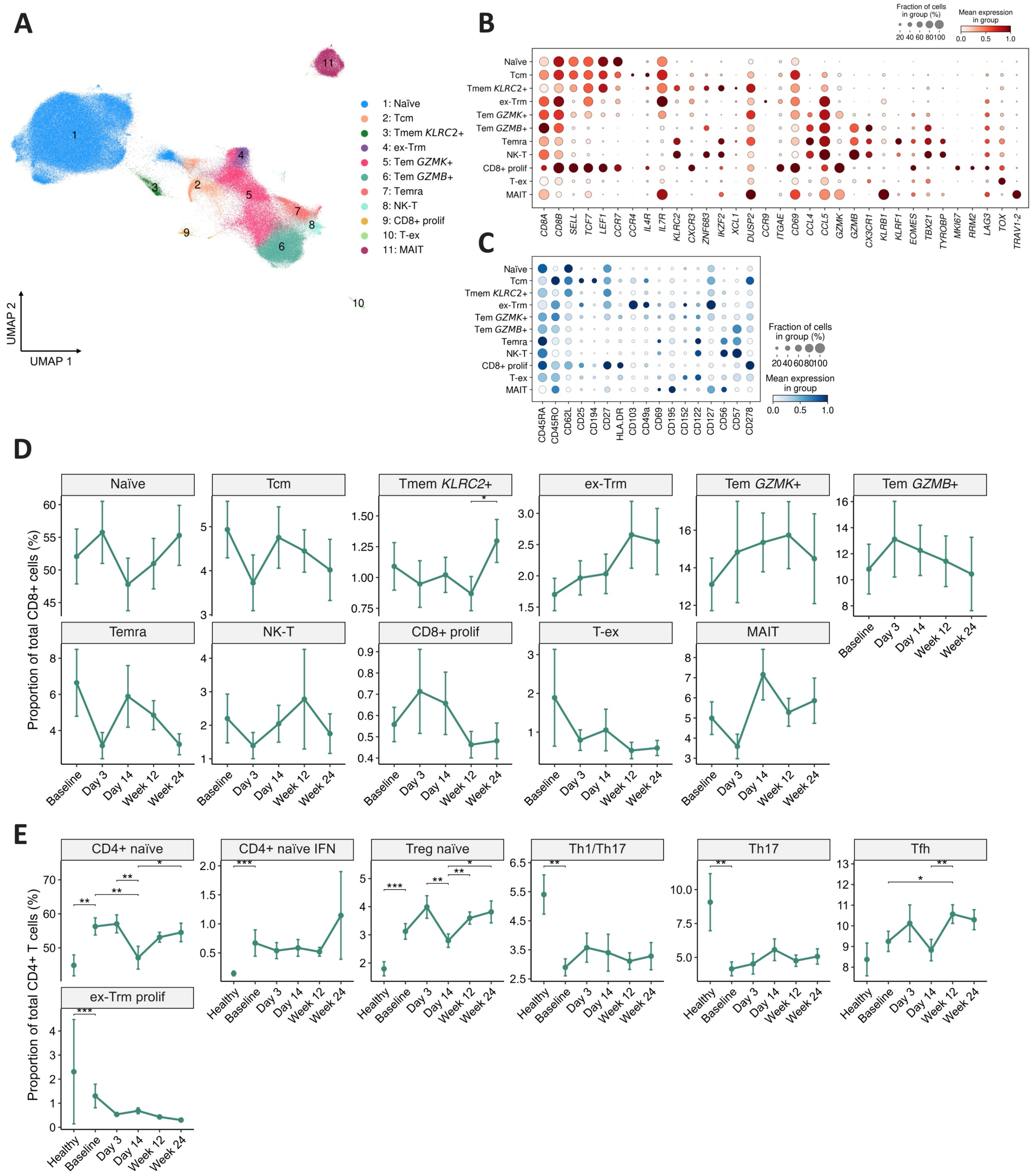


**Fig. S3**. **The abundance of CD8+ T and Th17 cells does not change significantly after IL4Rα blockade. A)** UMAP of 277,704 CD8+ T cells, identifying 13 subclusters. **B-C)** Dot plots showing the expression of RNA (B) and protein (C) markers used to annotate cell subsets. **D-E)** Line plots showing the abundance of CD8+ (D) and CD4+ (E) T cell clusters at various timepoints after treatment. Data are presented as mean +/- SEM. *$P$<0.05; **$P$<0.01; ***$P$<0.001 (mixed design ANOVA for time-point analysis; Wilcoxon rank-sum test for comparison between healthy controls and baseline AD, see Methods for further details). CD8+ T prolif, proliferating CD8+ T cells. Ex-Trm, ex-tissue resident memory T cells; MAIT, mucosal associated invariant T cells; TEMRA, Terminally Differentiated Effector Memory T cells; T-ex, exhausted T cells.

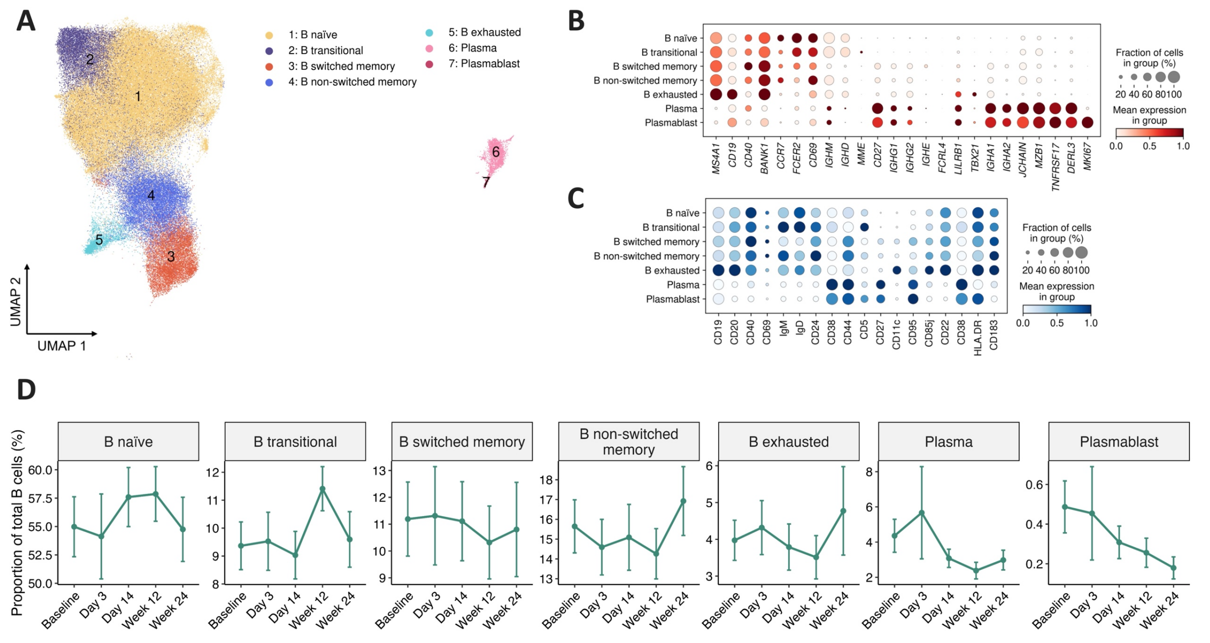


**Fig. S4**. **The abundance of B cells does not change significantly after IL4Rα blockade. A)** UMAP of 113,494 B cells clustered into seven populations. **B-C)** Dot plots showing the expression of RNA (B) and protein (C) markers used to annotate B cell subsets. **D)** Line plots showing cell cluster abundance at various timepoints after treatment. Data are presented as mean +/- SEM.

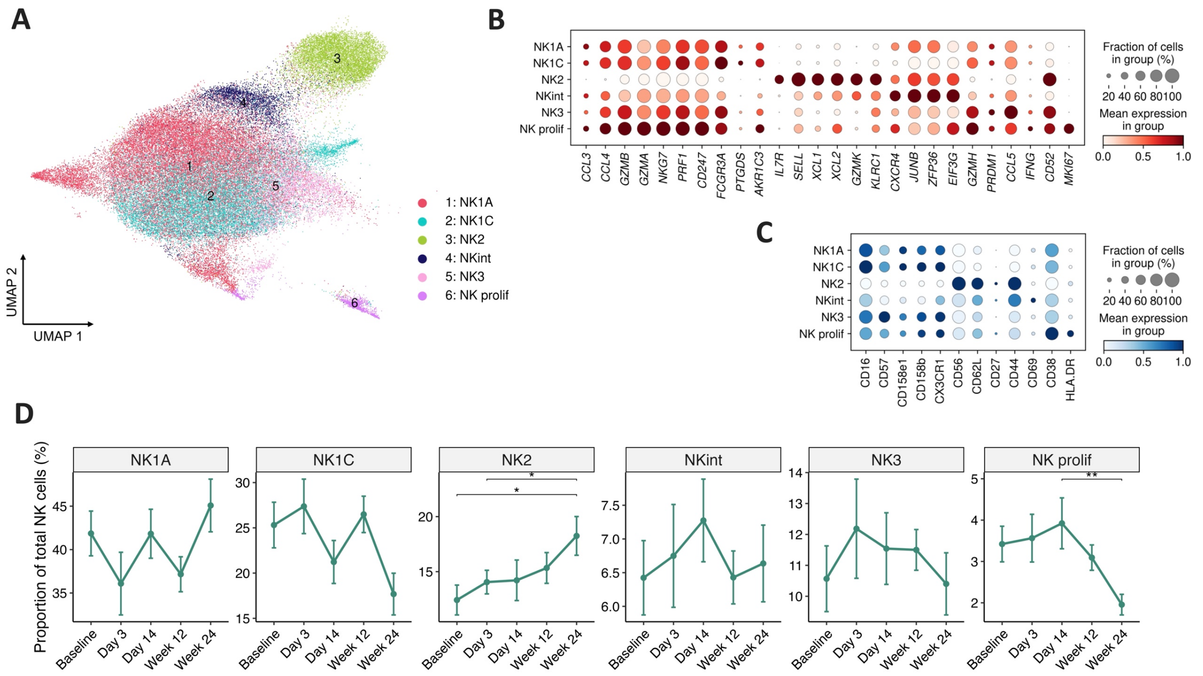


**Fig. S5. The abundance of natural killer (NK) cells does not change significantly after IL4Rα blockade. A)** UMAP of 62,947 NK cells, identifying six subclusters. **B-C)** Dot plots showing the expression of RNA (B) and protein (C) markers used to annotate NK cell subsets. **D)** Line plots showing cell cluster abundance at various timepoints after treatment. Data are presented as mean +/- SEM. **P*<0.05; ***P*<0.01 (mixed design ANOVA; post-hoc test: pairwise comparison of estimated marginal means with Benjamini-Hochberg correction). NK prolif, proliferating natural killer cells.

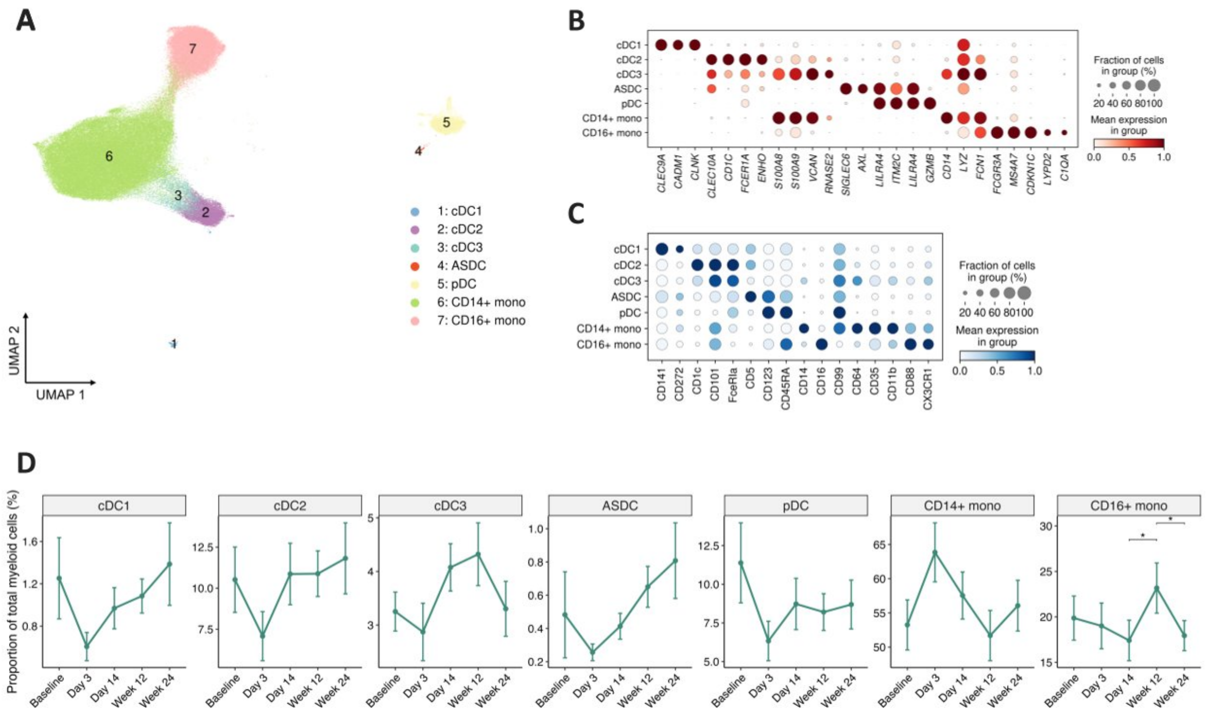


**Fig. S6**. **The abundance of circulating monocytes, myeloid dendritic cells or plasmacytoid dendritic cells does not change significantly after IL4Rα blockade A)** UMAP of 137,569 myeloid or plasmacytoid cells, identifying 8 subclusters. **B-C)** Dot plots showing the expression of RNA (B) and protein (C) markers used to annotate cell subsets. **D)** Line plots showing cell cluster abundance at various timepoints after treatment. Data are presented as mean +/- SEM. **P*<0.05 (mixed design ANOVA; post-hoc test: pairwise comparison of estimated marginal means with Benjamini-Hochberg correction). ASDC, *AXL+*, *SIGLEC6+* dendritic cells; cDC, conventional dendritic cells; mono, monocytes; pDC, plasmacytoid dendritic cells.

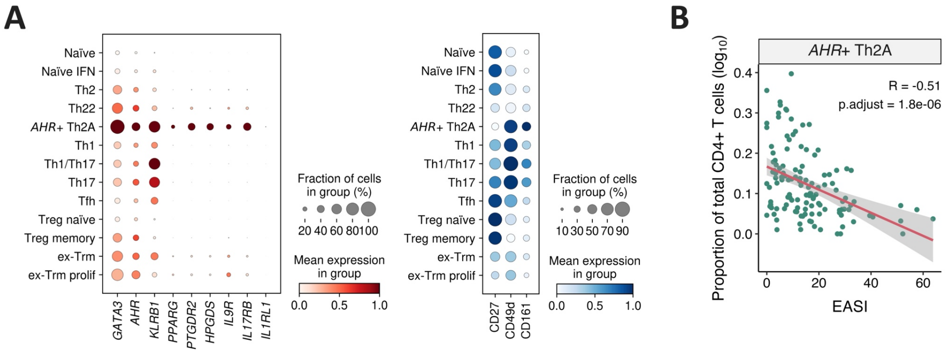


**Fig. S7.** ***AHR*+ Th2A cells have an immune regulatory profile. A)** Dot plots of CITE-seq RNA (left) and protein (right) readouts, showing the expression of markers used to annotate *AHR*+ Th2A cells. **B)** Inverse correlation between Eczema Area and Severity Index (EASI) and the abundance of *AHR*+ Th2A cells. Each dot is a sample; the regression line is shown in red with 95% confidence intervals shaded in grey. The p-value was calculated by repeated measure correlation analysis and adjusted using the Benjamini-Hochberg correction.

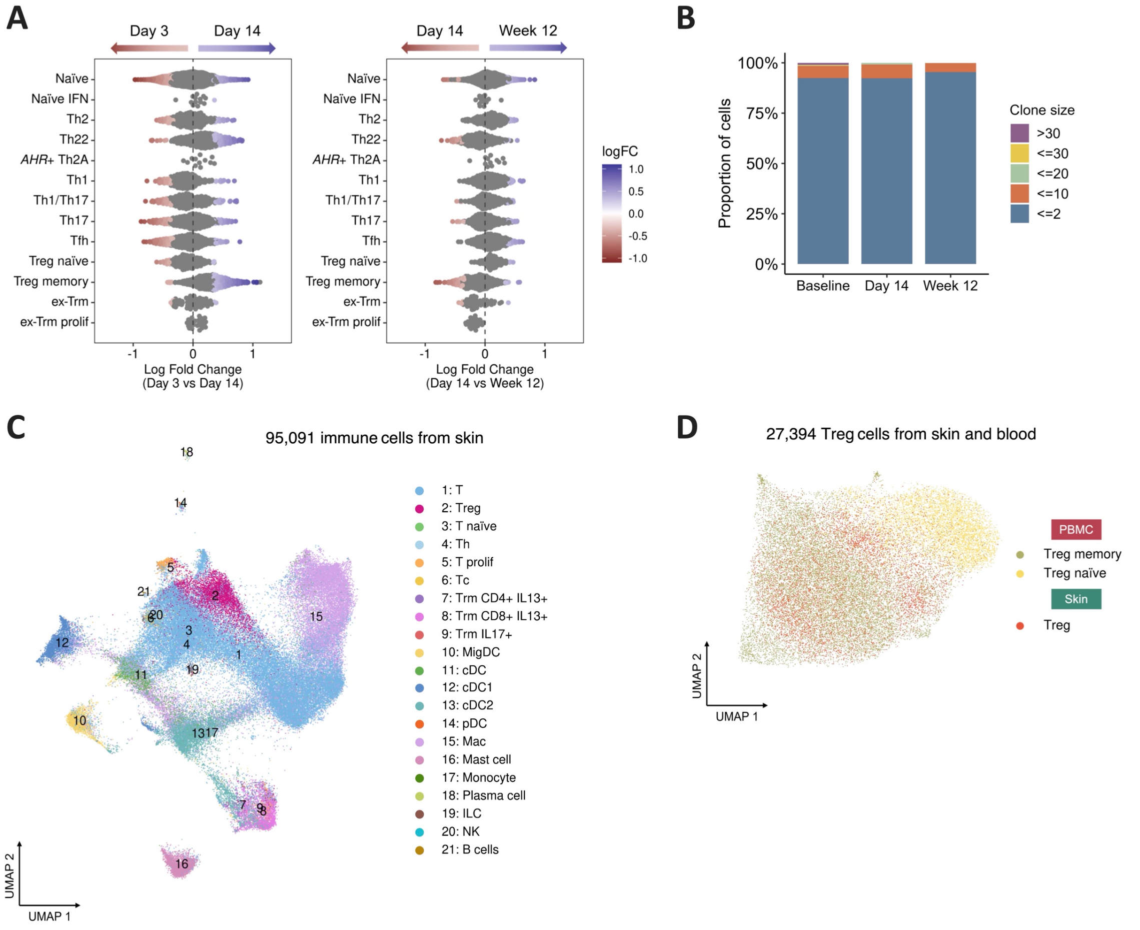


**Fig. S8**. **Changes in memory Treg cells during drug-induced remission. A)** Milo beeswarm plot: the memory Treg cells neighbourhoods that show differential abundance across time points are coloured in red or blue. **B)** Stacked bar plots showing Treg clonal expansion across time points. **C)** UMAP visualisation of 95,091 cells coloured based on their annotation. **D)** Integration of skin-resident and circulating Tregs, visualised as a UMAP.

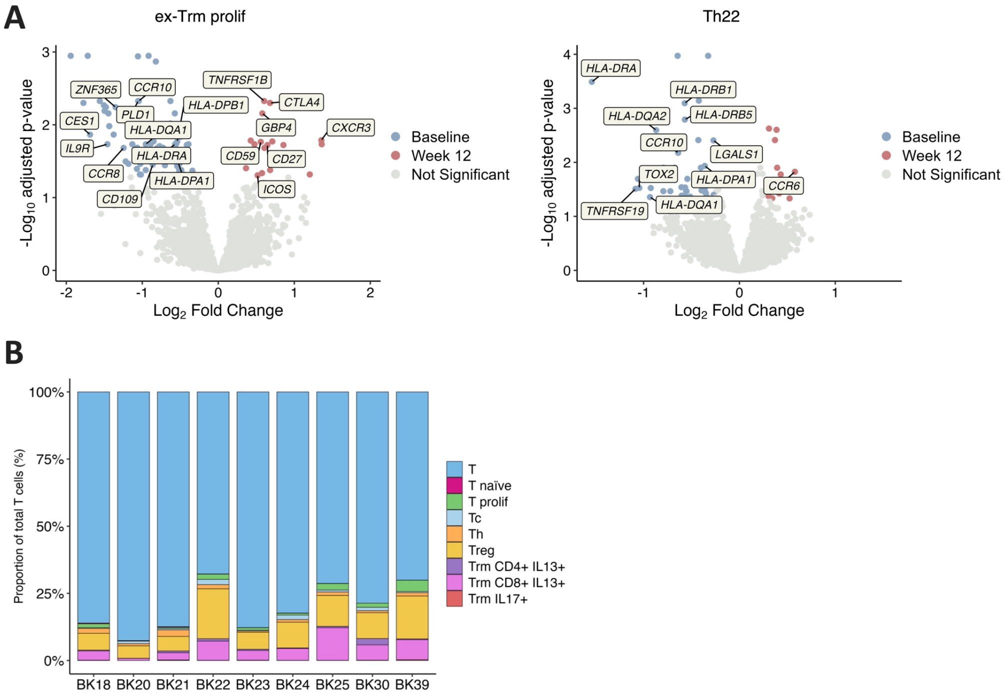


**Fig. S9**. **Drug-induced changes in immune populations related to CD4+ ex-Trm cells. A)** Differential expression analysis of proliferating ex-Trm (left) and Th22 (right) cells. The volcano plots show the genes that are differentially expressed (log2(fold change)>|log2(1.2), *FDR*<0.05) after 12 weeks of treatment. Coloured dots represent genes that are upregulated at baseline (blue) or week 12 (red). Labels show selected inflammatory genes. **B)** Stacked bar plots showing the abundance of resident T cell subsets in the skin of each donor.

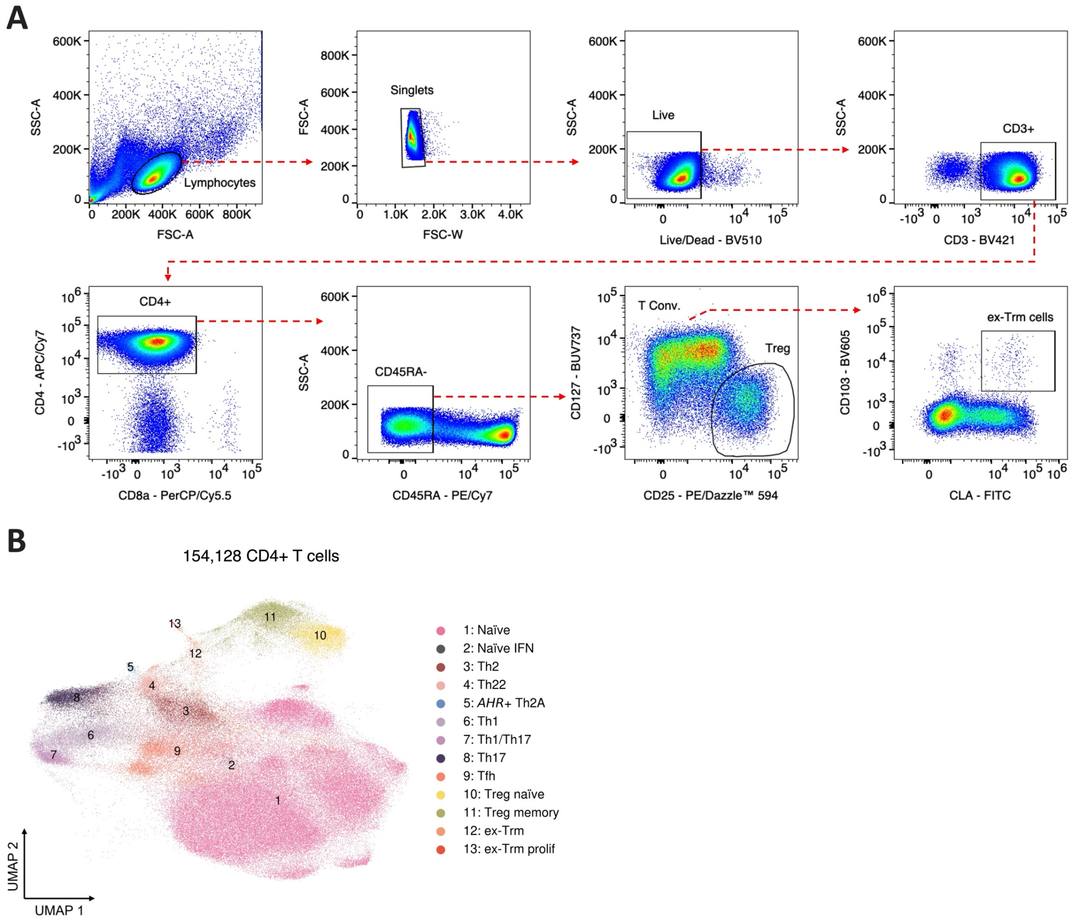


**Fig. S10**. **Analysis of circulating CD4+ ex-Trm cells in two validation cohorts. A)** Gating strategy for flow cytometry experiments. After removing doublets and non-viable cells, memory T-cells were identified as CD3+/CD4+/CD45RA- cells. Treg (CD25+CD127-) cells were removed from the analysis and the remaining cells were gated for CD103 and CLA expression. CD4+ ex-Trm cells were identified as a CD103+/CLA+ population. T Conv, conventional T cells (non-Treg memory T cells). **B)** UMAP visualisation of 154,128 PBMCs obtained from the 8 donors who discontinued dupilumab treatment after achieving durable AD remission (clear skin for >1 year).

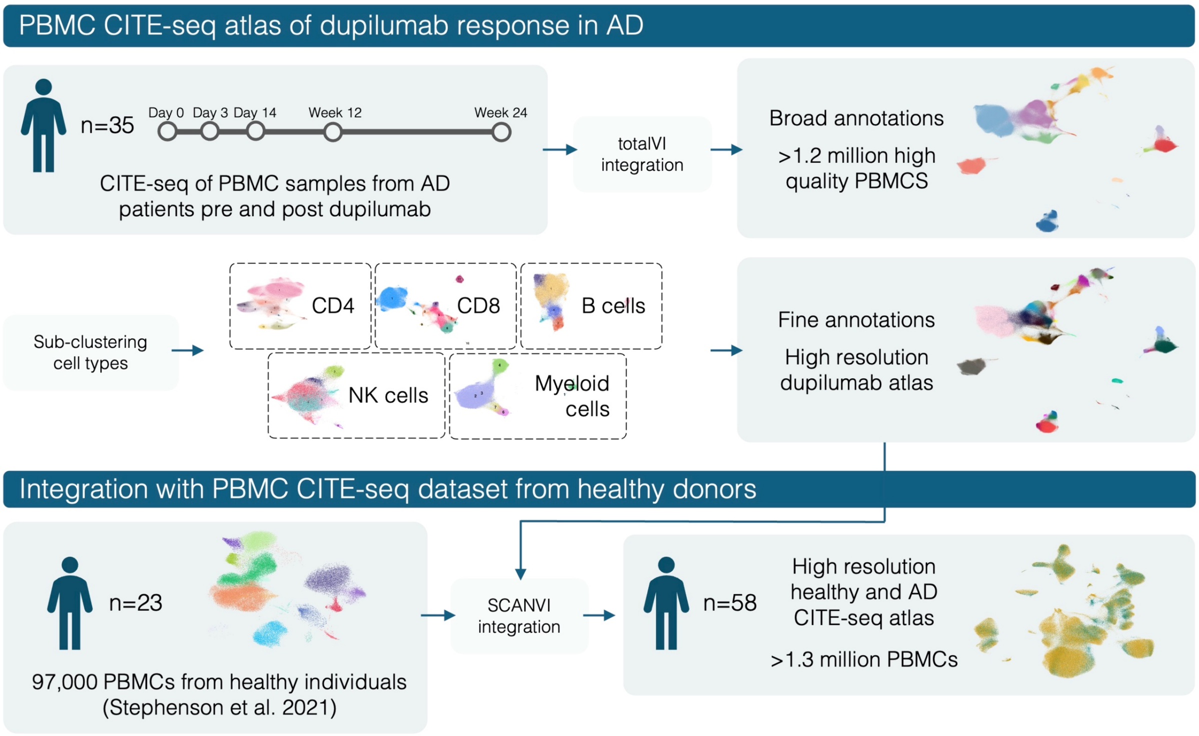


**Fig. S11**. **Data integration workflow.** The schematic illustrates how the longitudinal CITE-seq data generated in AD cases was integrated with a public resource, generated in healthy controls.

**Table S1**: Characteristics of the main study cohort

| | |
|---|---|
| *Recruitment* [n (%)] | |
| Routine clinical care | 31 (88.6%) |
| BEACON trial | 4 (11.4%) |
| *Demographics* | |
| Age [years, mean (SD)] | 39.1 (14.5) |
| Sex [n (%)] | |
| Male | 20 (57.1%) |
| Female | 15 (42.9%) |
| Ancestry [n, (%)] | |
| European | 25 (71.4%) |
| Asian | 5 (14.3%) |
| African-Caribbean | 2 (5.7%) |
| Unknown | 3 (8.6%) |
| *AD clinical features* [mean (SD)] | |
| Baseline EASI | 23.1 (13.0) |
| Itch score | 6.5 (2.0) |
| Age of onset (years) | 4.8 (12.9) |
| *Comorbidities* [n (%)] | |
| Allergy | 28 (80.0%) |
| Asthma | 27 (77.1%) |
| Obesity[1] | 9 (25.7%) |
| *Prior AD treatment* [n (%)] | |
| Methotrexate | 17 (48.6%) |
| Ciclosporin | 13 (37.1%) |
| Azathioprine | 9 (25.7%) |
| Mycophenolate mofetil | 6 (17.1%) |
| Tralokinumab | 1 (2.9%) |
| *Clinical response to dupilumab* | |
| EASI-50 at week 24 [n (%)] | 35 (100%) |

[1]Body Mass Index ≥30kg/m$^2$; EASI, eczema area and severity index; EASI-50, 50% reduction in EASI.

**Table S2**: Characteristics of the flow cytometry validation cohort

| | |
|---|---|
| *Recruitment* [n (%)] | |
| Routine clinical care | 6 (66.7%) |
| BEACON trial | 3 (33.3%) |
| *Demographics* | |
| Age [mean (SD)] | 34.7 (9.0) |
| Sex [n (%)] | |
| Male | 5 (55.6%) |
| Female | 4 (44.4%) |
| Ancestry [n, (%)] | |
| European | 5 (55.6%) |
| Asian | 3 (33.3%) |
| Mixed | 1 (11.1%) |
| *Disease Presentation* [mean (SD)] | |
| Baseline EASI | 26.7 (8.6) |
| Age of onset | 2.6 (4.0) |
| *Comorbidities* [n (%)] | |
| Allergy | 9 (100%) |
| Asthma | 3 (33.3%) |
| Obesity[1] | 1 (11.1%) |
| *Prior AD treatment* [n (%)] | |
| Methotrexate | 5 (55.6%) |
| Ciclosporin | 2 (22.2%) |
| Azathioprine | 1 (11.1%) |
| *Response to Treatment* [n (%)] | |
| EASI-50 at week 24 | 7[2] (100%) |

[1]Body Mass Index ≥30kg/m$^2$; [2]One patient is not yet due for week 24 visit, another patient did not attend.

**Table S3**: Characteristics of the drug withdrawal cohort

| | |
|---|---|
| *Recruitment* [n (%)] | |
| Routine clinical care | 8 (100%) |
| *Demographics* | |
| Age [years, mean (SD)] | 44 (12.2) |
| Sex [n (%)] | |
| Male | 3 (37.5%) |
| Female | 5 (62.5%) |
| Ancestry [n, (%)] | |
| European | 4 (50%) |
| Asian | 3 (37.5%) |
| Other | 1 (12.5%) |
| *Comorbidities* [n (%)] | |
| Allergy | 6 (75%) |
| Asthma | 6 (75%) |
| Obesity[1] | 3 (37.5%) |
| *AD disease course [mean (SD)]* | |
| Age of onset (years) | 2.9 (3.9) |
| EASI score at withdrawal | 0.15 (0.3) |
| Time to recurrence[2] (weeks) | 6.4 (2.0) |

[1]Body Mass Index $\geq$30kg/m$^2$, [2]After drug withdrawal.

**Table S4**: Flow cytometry antibodies

| *Antibody details* | *Supplier* | *Dilution* |
|---|---|---|
| Monoclonal Mouse Anti-Human CD127-BUV737 [Clone: HIL-7R-M21] | BD Biosciences | 1:50 |
| Monoclonal Mouse Anti-Human CD3-BV421 [Clone: UCHT1] | BioLegend | 1:100 |
| Monoclonal Mouse Anti-Human CD103-BV605 [Clone: Ber-ACT8] | BioLegend | 1:50 |
| Monoclonal Rat Anti-Human/Mouse CLA-FITC [Clone: HECA-452] | BioLegend | 1:100 |
| Monoclonal Mouse Anti-Human CD8a-PerCP/Cyanine5.5 [Clone: RPA-T8] | BioLegend | 1:100 |
| Monoclonal Mouse Anti-Human CD25-PE/Dazzle™ 594 [Clone: M-A251] | BioLegend | 1:100 |
| Monoclonal Mouse Anti-Human CD45RA-PE/Cyanine7 [Clone: HI100] | BioLegend | 1:1000 |
| Monoclonal Mouse Anti-Human CD4-APC/Cyanine7 [Clone: RPA-T4] | BioLegend | 1:400 |
